# Nationwide SARS-CoV-2 Seroprevalence Trends in the Netherlands in the Variant of Concern Era, 2021-2022: an Ongoing Prospective Cohort Study

**DOI:** 10.1101/2023.11.22.23298889

**Authors:** Eric R.A. Vos, Cheyenne C.E. van Hagen, Denise Wong, Gaby Smits, Marjan Kuijer, Alienke J. Wijmenga-Monsuur, Joanna Kaczorowska, Robert S. van Binnendijk, Fiona R.M. van der Klis, Gerco den Hartog, Hester E. de Melker

## Abstract

**Background:** Repeated population-based SARS-CoV-2 serosurveillance is key in complementing other surveillance tools.

**Aim:** Assessing trends in infection- and/or vaccine-induced immunity, including breakthrough infections, among (sub)groups and regions in the Dutch population during the Variant of Concern (VOC)-era whilst varying levels of stringency, to evaluate population immunity dynamics and inform future pandemic response planning.

**Methods:** In this prospective population-based cohort, randomly-selected participants (n=9,985) aged 1-92 years (recruited since early-2020) donated home-collected fingerstick blood samples at six timepoints in 2021-2022, covering waves dominated by Alpha, Delta, and Omicron (BA.1, BA.2, BA.5). IgG antibody assessments against Spike-S1 and Nucleoprotein were combined with vaccination- and testing data to estimate infection-induced (inf) and total (infection- and vaccination-induced) seroprevalence.

**Results:** In 2021, nationwide inf-seroprevalence rose modestly from 12% since Alpha to 26% amidst Delta, while total seroprevalence increased rapidly to nearly 90%, particularly fast in vulnerable groups (i.e., elderly and those with comorbidities). Highest infection rates were noticeable in adolescents and young adults, low/middle educated elderly, non-Western, contact professions (other than healthcare), and low-vaccination coverage regions. In 2022, following Omicron emergence, inf-seroprevalence elevated sharply to 62% and further to 86%, with frequent breakthrough infections and reduction of seroprevalence dissimilarities between most groups. Whereas >90% of <60-year-olds had been infected, 30% of vaccinated vulnerable individuals had not acquired hybrid immunity.

**Conclusion:** Although total SARS-CoV-2 seroprevalence had increased rapidly, infection rates were unequally distributed within the Dutch population. Ongoing tailored vaccination efforts and (sero-)monitoring of vulnerable groups remain important given their lowest rate of hybrid immunity and highest susceptibility to severe disease.

## INTRODUCTION

Severe acute respiratory syndrome coronavirus-2 (SARS-CoV-2), causative agent of coronavirus disease 2019 (COVID-19), has imposed a tremendous burden on societies worldwide since its origin in late 2019. In the Netherlands, stringent measures including social distancing and several lockdowns (with fluctuating stringency (**Figure 1A**)) have been initiated to curb transmission and prevent the health system from collapsing. Nonetheless, initial waves of infection in spring 2020 and later in winter caused huge spikes in hospitalizations and over 20,000 deaths [1]. Although some regions were hit relatively hard, only a small proportion of the population had been infected [2, 3]. A nationwide vaccination campaign started in January 2021 and followed an age-, comorbidity- and healthcare worker (HCW)-based prioritization [4]. Vaccination coverage was generally high and effectively changed the epidemiology, yet at the same time Variants of Concern (VOC) emerged. The increased transmissibility [5] and severity of Alpha and, thereafter, Delta caused a steep rise in reported cases, hospital- and ICU admissions and deaths predominantly among unvaccinated individuals [5-10], thus mainly affecting groups with low access and uptake of vaccines [11]. The increasing number of reports of waning vaccine effectiveness triggered administration of booster doses in autumn 2021 [12]. Signs of enhanced immune escape became even more apparent with emergence of Omicron in late 2021, reflected by high numbers of reported (breakthrough) cases. Relatively fewer persons became severely-ill, enabling relaxation of control measures in spring 2022 [10, 13].

**Figure 1.**
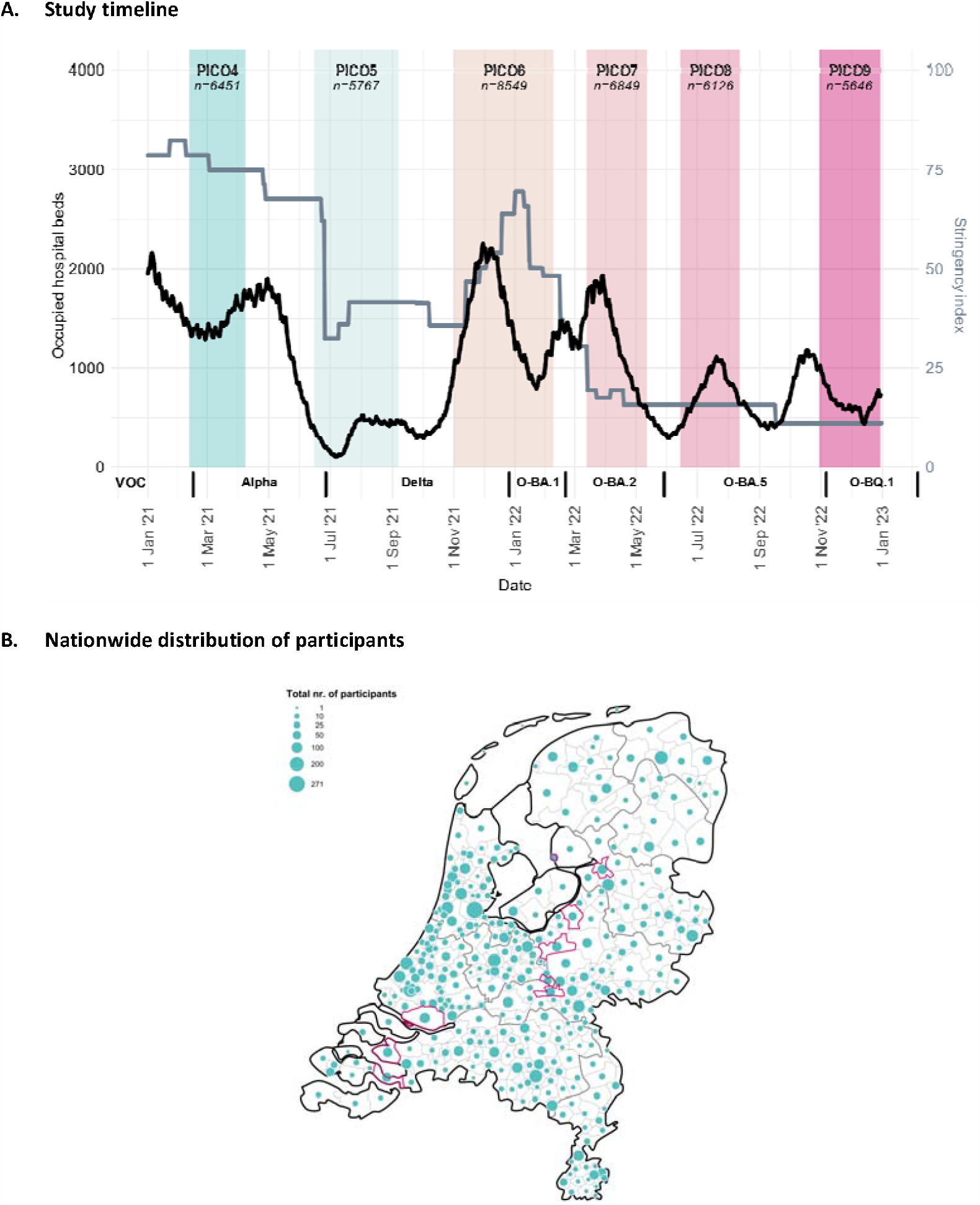
A. The PIENTER Corona (PICO) study timeline in 2021 and 2022, by daily occupied hospital beds due to COVID-19 (black line, left y-axis), daily stringency index (grey line, right y-axis) and dominance of specific Variants of Concern (VOC) in the Netherlands (on x-axis, from left to right: Alpha, Delta, Omicron-BA.1, -BA.2, -BA.5, and -BQ.1). A nationwide vaccination campaign started in the beginning of January 2021 and followed an age-, comorbidity- and healthcare worker-based prioritization. The number of participants analyzed in the current study are provided per study round and depicted in italic in the colored boxes. Length of the study rounds are consistent with the width of the colored boxes, although the majority of participants participated at the beginning of each study round, also reflected by the median and interquartile ranges (IQR): PICO4: 17 (15-19) February 2021; PICO5: 23 (21-28) June 2021; PICO6: 11 (8-15) November 2021; PICO7: 23 (19-28) March 2022; PICO8: 21 (19-27) June 2022; and PICO9: 9 (5-14) November 2022. Data on hospital bed occupation are open source and were downloaded from the National Coordination Center for Patient Distribution (LCPS) (via: https://lcps.nu/datafeed). Stringency index data, here depicted for the Netherlands, are open source too and were downloaded from Our World in Data (via: https://ourworldindata.org/covid-stringency-index. **B. Nationwide distribution of the total number of unique PICO participants analyzed (n=9,985), by municipality.** The current study comprises the national sample (n=9,492) and the low vaccination coverage (LVC) sample (n=493). The number (proportional to the size of the green dots) and distribution of participants match the countrywide population size and -distribution. Pink-circled municipalities represent the LVC municipalities (as per the PIENTER-3 cohort). Thicker grey lines represent the borders of the provinces and the thin grey lines those of the municipalities.

To assist public health decision-making during different phases of the pandemic, e.g., regarding effectiveness of control measures and vaccination, or insights on severity of disease, it is essential to understand population dynamics of infection and vaccination, and potential disparities between groups [3, 8]. However, traditional surveillance methods underestimate prevalence and incidence as they rely on testing policy and (self/home) testing behavior, or because a proportion of cases is asymptomatic and remains undiagnosed [14]. Hence, seroepidemiology provides a unique opportunity in complementing other surveillance tools by estimating the prevalence of antibodies induced by infection and/or vaccination, generally indicative of protection against (severe) disease. However, the majority of serosurveys have a number of limitations, for instance: (1) usage of convenience samples that lack representativeness of the general population and in-depth questionnaire data, such as sample sets consisting of only healthcare workers or blood donors, or not including children and elderly; (2) usage of cross-sectional sample sets which carry a risk of missing previous infections due to waning immunity; or (3) sole qualitative assessment of the presence of antibodies, which limits the detection of re-infections [15].

Therefore, by means of the large nationwide population-based prospective PIENTER Corona (PICO) serosurveillance study, we sero-monitored the Dutch population across all ages throughout the acute phase of the pandemic in order to inform (short-term) decision-making. Here, we aim to assess trends in infection- and/or vaccine-induced immunity, including breakthrough infections, among different (sub)groups and regions in the population during the VOC-era in 2021-2022. These findings can serve as input for evaluation of the pandemic response and construction of future preparedness globally, and guide ongoing preventive strategies in the endemic/post-pandemic period.

## METHODS

### Study population and materials

Participants were recruited from the Dutch population registry via random selection in a region- and age-specific manner. Details on the sampling at the set-up of the PICO-study (using the framework of the PIENTER3 serosurvey established in 2016/17 [16]) and the additional sampling prior to round 2 (PICO2, June 2020, after the first wave) have been described comprehensively [2, 3]. The PICO-cohort consists of a national sample with participants from across the country resembling the Dutch population (NL); and a sample with participants from municipalities with a lower childhood vaccination coverage (LVC, conceived in PIENTER3), inhabited by a relatively large portion of Orthodox-Reformed Protestants who largely refuse vaccination on religious grounds (commonly known as ‘the Bible belt’). Prior to PICO6 (November 2021), NL was supplemented equal to previous sampling with randomly-selected persons (predominately younger and older age groups due to drop-outs) to maintain power. Details are provided in the **Supplementary information** and **Supplementary Table S1**.

In the current study, participants were followed-up at six timepoints during the VOC-era (2021-2022), covering PICO4-9. Median inclusion date in PICO4 (17 Feb 2021) matched the start of Alpha dominance (i.e., PICO4 findings display the pre-Alpha period). Intervals between median dates of subsequent rounds greatly match waves dominated by single VOCs (except for Delta that covered PICO6 and also partly PICO7 due to its dominance up until the end of Dec 2021), namely: Alpha (PICO5: 23 June 2021), Delta (PICO6: 11 Nov 2021), Omicron-BA.1 (PICO7: 23 Mar 2022), Omicron-BA.2 (PICO8: 21 Jun 2022), and Omicron-BA.5 (PICO9: 9 Nov 2022) (**Figure 1A**). Each round, fingerstick blood samples were home-collected using contact-activated lancets (BD microtainer) and mini-collect serum tubes (Greiner) which were returned to the RIVM-laboratory in safety envelopes. Serum was processed and stored at −20°C awaiting analyses. Concurrently, participants were requested to complete a(n) (online) questionnaire regarding sociodemographics, occupation, comorbidities, self-reported SARS-CoV-2 testing (PCR/rapid-antigen test) and vaccination data, and factors related to potential exposure.

All persons aged ≥12 years had been offered primary vaccination before the end of 2021. Since autumn 2021, booster vaccination was provided (firstly) to vulnerable groups and HCW, and later to everyone ≥18 years during winter [4]. Since the beginning of 2022, primary vaccination has been available for 5-11 years-olds, yet nationwide coverage has been low (2%) [10]. Second boosters, targeted to at-risk groups, were offered in spring of 2022, and bivalent boosters followed that autumn [17].

### Serological analyses

Serum samples were quantitatively analysed for SARS-CoV-2 immunoglobulin G (IgG) antibodies against Spike-S1 and Nucleoprotein (N) antigens using a validated fluorescent bead-based multiplex-immunoassay, as described previously [18]. Briefly, samples were diluted and incubated with S1-(Sino Biological, 40591-V08H) and N- (Sino Biological, 40588-V08B) coupled beads in SM01-buffer (Surmodics, USA) supplemented with 2% FCS, while shaking at room temperature in the dark for 45 minutes. Plates were washed three times with PBS, and incubated with PE-conjugated goat anti-human IgG (Jackson ImmunoResearch, 109-116-098) for thirty minutes. Following final washing steps, samples were acquired on a Luminex FlexMap3D. Antibody concentrations were interpolated from pooled sera calibrated against the World Health Organization (WHO) standard (NIBSC, 20/136) using a 5-parameter logistic fit. Seropositivity for anti-S1 was considered at a cut-off concentration of 10 binding antibody units (BAU) per milliliter (mL) and for anti-N at 14.3 BAU/mL, as derived from previous mixture modelling and receiver operator characteristics analyses [3, 19]. Applying these cut-offs to detect infection resulted in a specificity for anti-S1 of 99.9% and a sensitivity of 94.3%, and for anti-N of 98.0% and 86.0%, respectively.

### Statistical analyses

Data cleaning, management and analyses were performed in SAS v.9.4. (SAS Institute Inc., USA) and R v.4.2.2.

At each study round, serological information (since the start of the study) was combined with vaccination- and testing data to assess infection-induced (inf) (at least once) and total (i.e., infection- and vaccination-induced) seroprevalence (equivalent to S1-seropositivity). More specifically, inf-seroprevalence was determined via anti-S1 seropositivity in unvaccinated (who could potentially sero-revert); and in vaccinated individuals via anti-N seropositivity, a SARS-CoV-2 positive test, or a

≥4-fold-increase in anti-S1 given they had not received a vaccine dose four weeks before their previous collection (i.e., the time for a vaccine response to peak, thereby ruling-out a potential further increase since previous collection due to vaccination and thus fold-increase in the current round). Participants aged ≥12 years enrolled since PICO6 were excluded for these analyses to prevent underestimation of previous infection as anti-N wanes relatively fast (which had to be relied on since most were vaccinated) and they lacked serological history [19]. Moreover, breakthrough infections could be identified in vaccinated persons who presented a positive SARS-CoV-2 test (at least 14 days) after their first vaccination, seroconverted for anti-N in rounds after their first vaccination or had a ≥4-fold-increase if already anti-N seropositive, or had a ≥4-fold-increase in anti-S1 given they had not received a vaccine dose four weeks before their previous collection.

Whole population (overall) and (sub)groups seroprevalence estimates (total and inf), vaccination coverage and -doses, and proportion of breakthrough infections (and their 95% confidence intervals (CIs)) were calculated. The survey design was taking into account and weights (per study sample) were incorporated using a set of sociodemographic characteristics (age, sex, ethnic background and urbanization degree) to match the Dutch population distribution following census data (of 1 January 2020) from the Statistics Netherlands. Infection estimates were controlled for test performance characteristics subsequently [20]. Differences in seroprevalence between (sub)groups were determined by estimating the parameters of the beta distribution for these estimates using the methods of moments [21]. Risk ratios, their corresponding 95% CIs and p values were estimated by Monte Carlo simulations of the seroprevalence estimates. P values <0.05 were considered statistically significant. Smooth age-specific seroprevalence estimates were modelled with surveylogistic regressions incorporating B-splines, using Akaike’s information criteria (AIC) for optimization of the number of percentile-placed knots.

## RESULTS

### Study population

Table 1 shows the sociodemographic characteristics, comorbidity- and vaccination status of participants throughout the study period. The total number of unique participants with a serological assessment in the current cohort was 9,985 and fluctuated between 5,767-8,549 across study rounds; of which NL comprised 5,371-8,138 and LVC 275-470. Drop-outs ranged between 8-20% per study round, yet the proportions of all sociodemographics groups were comparable over time. In NL, age ranged between 1-92 years, with a median between 51-57 years, and generally highest participation in adults up to 80 years. The five sampled regions were consistently similarly (∼20%) distributed across the cohort and over time (**Supplementary Figure S1 & Figure 1B**). Participants were more often women (56-57%); of native Dutch origin (88-89%), as compared to Western (7-9%) and non-Western (3-4%); and of lower urbanization degree (49-50%), versus moderate (31-32%) or high (19-20%). Educational level (high vs. middle/low) was nearly equally divided each round. One third could be classified as having comorbidities in 2021, which slightly increased to four out of ten in 2022. The majority of LVC-participants were of native Dutch, low urbanization degree (both >97%), low/middle educational level (71-73%), and median age as well as the proportion of having comorbidities (18-24%) was lower than in NL.

**Table 1.**
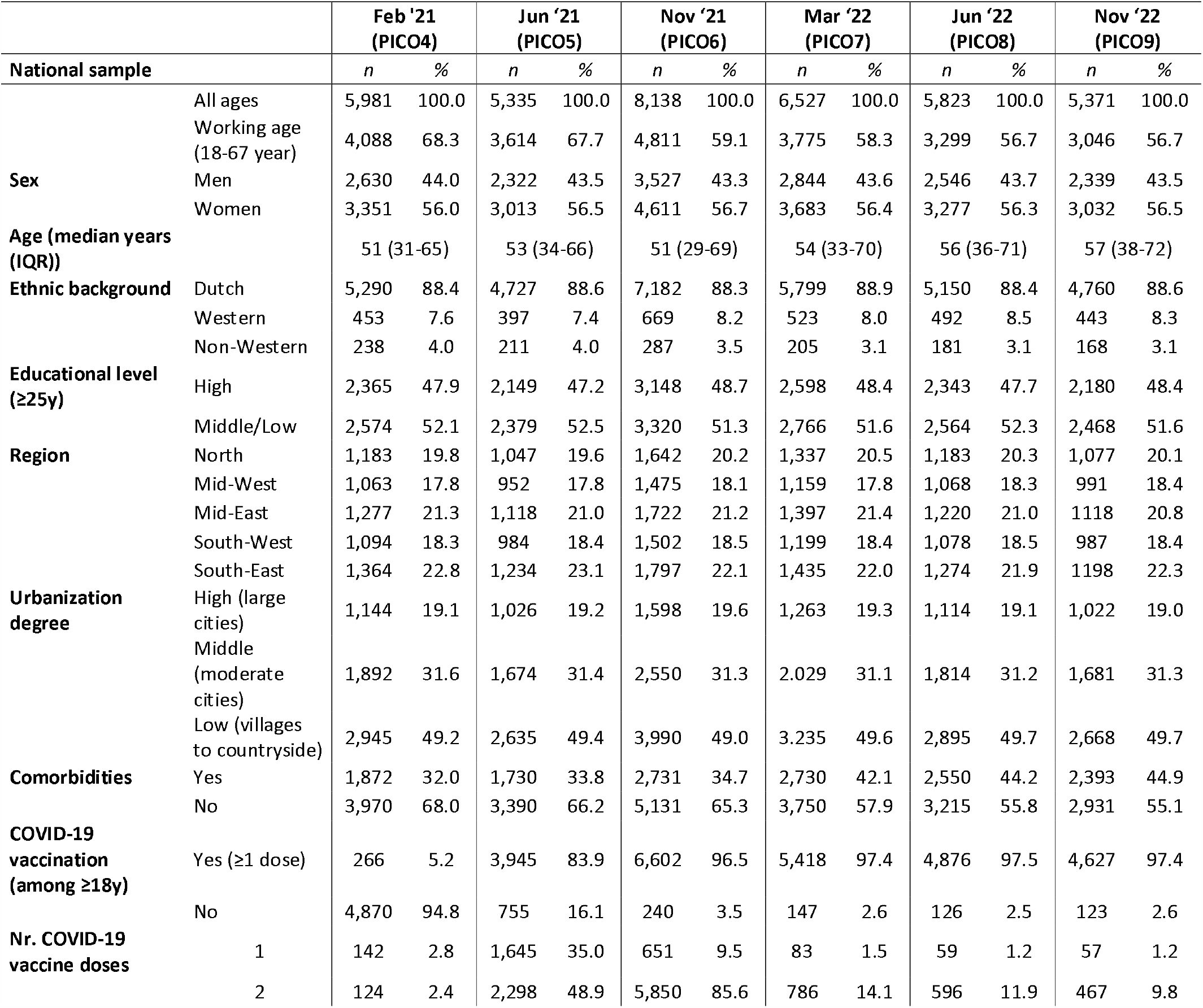
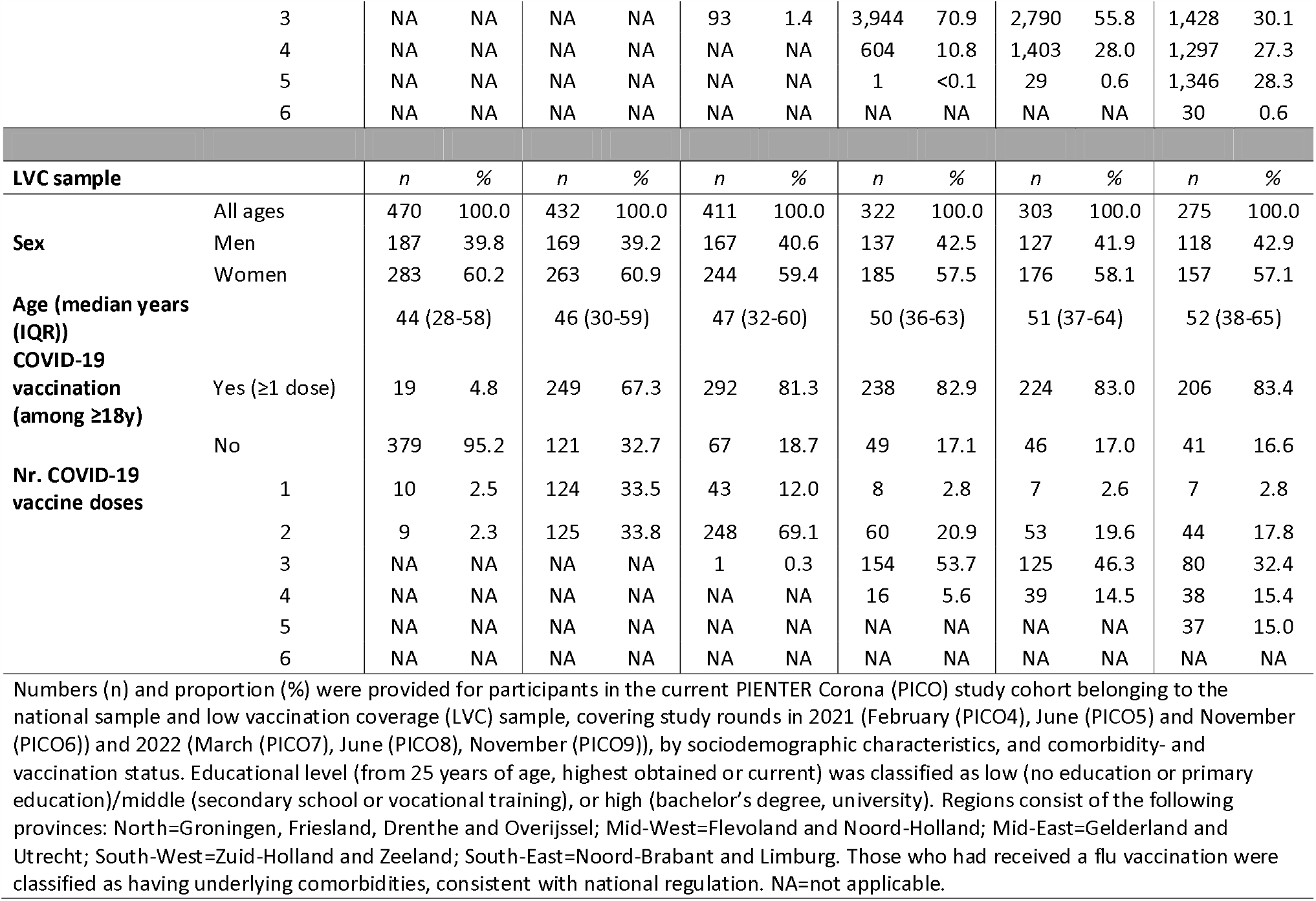
Description of the PIENTER Corona (PICO) study cohort in 2021 and 2022, covering study rounds 4 to 9.

COVID-19 vaccination coverage as well as the number of doses followed a distinct age-pattern over time in NL, consistent with the vaccine roll-out (**Supplementary Figure S2**). Overall weighted vaccination coverage (≥1 dose) in ≥18 years was 5% in February 2021, rose steeply to 78% in June, and increased further to 96% in November 2021. In March 2022, 78% had received a booster dose, and in June 18% had been administered a second booster. The latter increased to 42% in November, while 17% had received a fifth dose. Overall coverage did not differ much between sexes, yet was slightly higher in women <60 years in June 2021 for the primary series, but this difference was nearly erased by the end of the year and coverage remained stable for the doses thereafter. Uptake was consistently lower in the LVC (**Table 1**).

### Infection-induced and total seroprevalence

#### Overall, by age and region

The overall and stratified, inf- and total seroprevalence are displayed in **Figure 2** for 2021 (**A**) and 2022 (**B**), and by age in **Figure 3** (**A**: inf; **B**: total).

**Figure 2.**
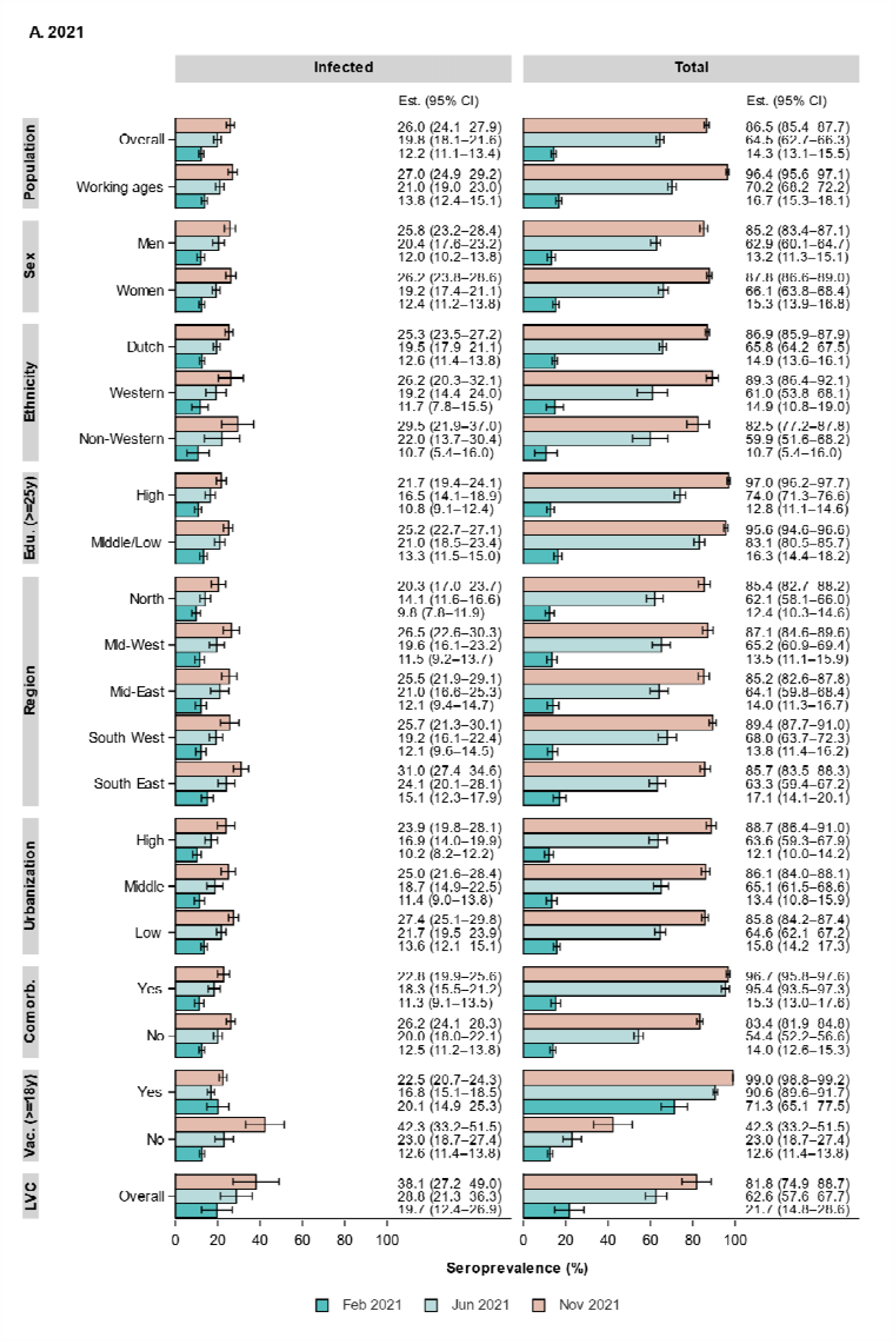
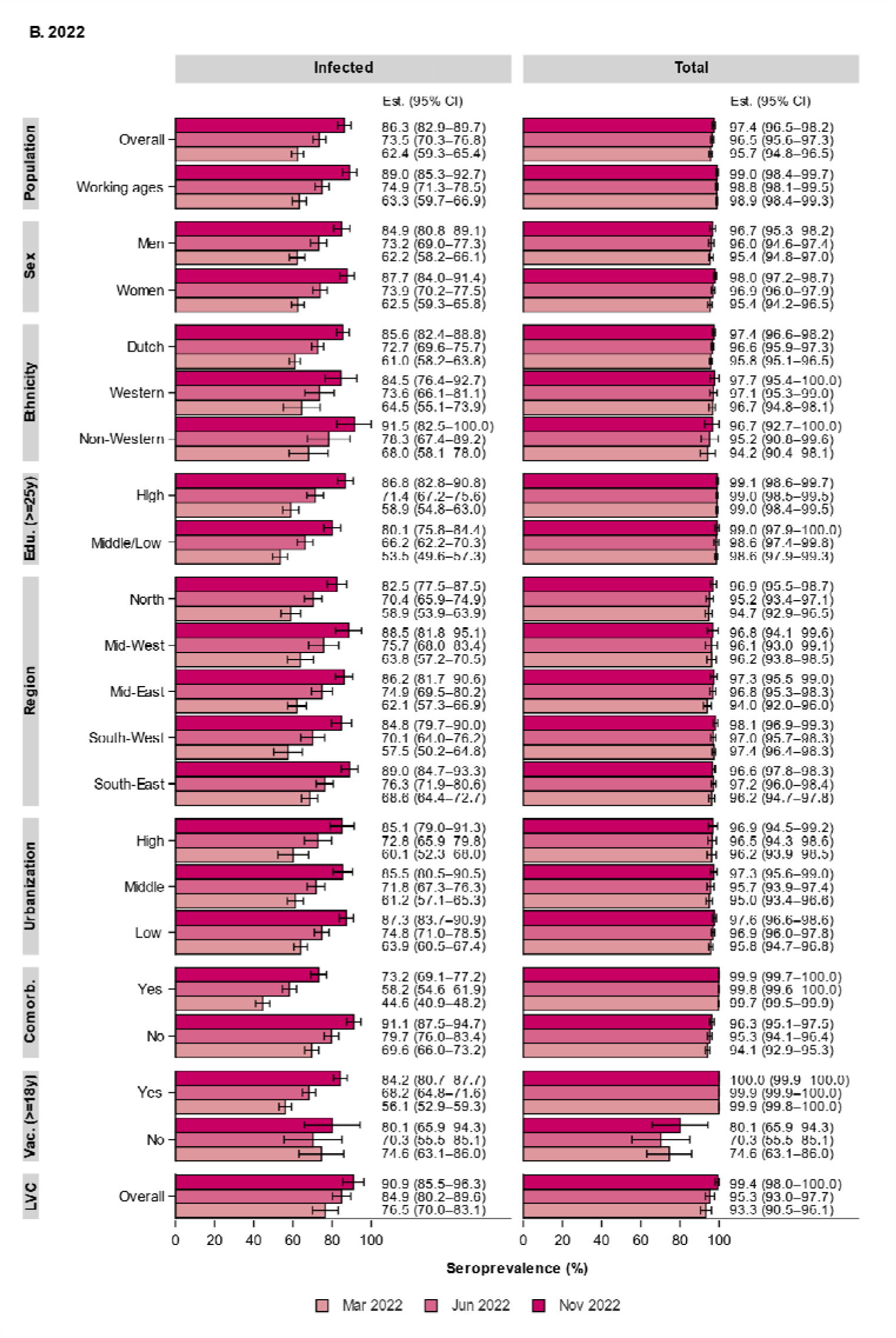
Weighted SARS-CoV-2 seroprevalence (with 95% confidence intervals (CI)) induced by infection (left panel) and total (i.e., infection and vaccination, right panel) in the general Dutch population in 2021 (February (PICO4), June (PICO5) and November (PICO6)) (A) and 2022 (March (PICO7), June (PICO8), November (PICO9)) (B), by sociodemographic characteristics, comorbidity-, and vaccination (>=18 years) status. Regions consist of the following provinces: North=Groningen, Friesland, Drenthe and Overijssel; Mid-West=Flevoland and Noord-Holland; Mid-East=Gelderland and Utrecht; South-West=Zuid-Holland and Zeeland; South-East=Noord-Brabant and Limburg. Comorb.=having comorbidities, i.e., classification based on having received the annual flu vaccination, consistent with national regulation; Edu.=educational level (from 25 years of age, highest obtained or current), classified as low (no education or primary education)/middle (secondary school or vocational training), or high (bachelor’s degree, university); Est. (95% CI)=seroprevalence estimate with 95% confidence intervals; LVC=low vaccination coverage sample, as conceived in the PIENTER-3 cohort); Population=general Dutch population, stratified by all ages (overall) and working ages (18 to 67 years); Vac.=had a COVID-19 vaccination at least once (from 18 years of age).

**Figure 3.**
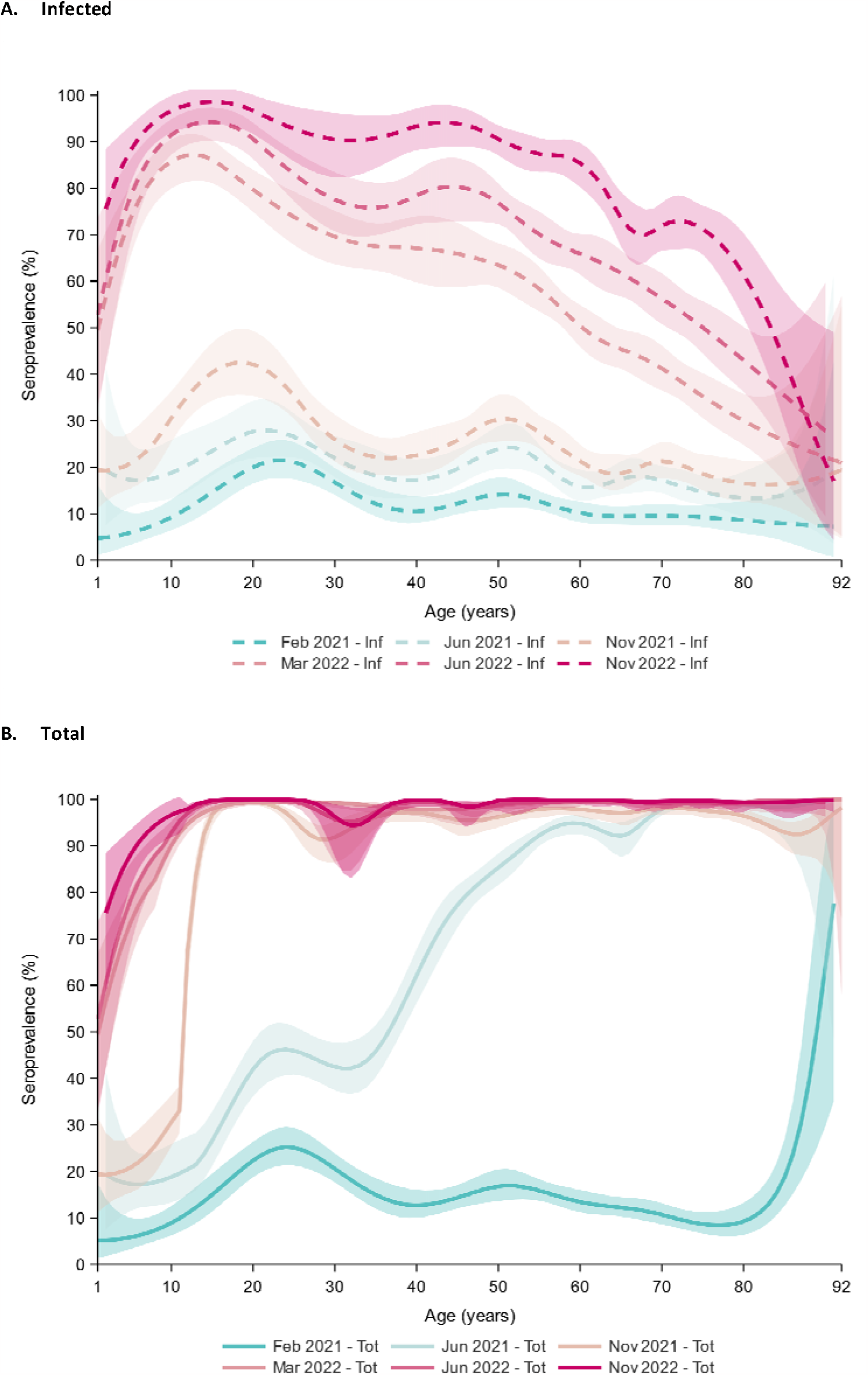
Weighted SARS-CoV-2 seroprevalence (with 95% confidence bands) induced by infection (A) and total (i.e., infection and vaccination) (B) in the general Dutch population in 2021 (February (PICO4), June (PICO5) and November (PICO6)) and 2022 (March (PICO7), June (PICO8), November (PICO9)), by age. Seroprevalence was modeled with B splines, with number of percentile-placed knots optimized following Akaike’s information criterion (AIC)).

In February 2021, pre-Alpha, the total seroprevalence reached 14% in the Dutch population and already spiked in the oldest ages, predominantly due to primary series vaccination. This continued with rapid pace during Alpha, rising overall to 65% in June, and among ≥50-years-olds to >80%. Meanwhile, overall inf-seroprevalence rose from 12% to 20%, and displayed a sustained age-pattern with highest estimates in young adults (30%), followed by middle-aged adults (25%), and lowest in elderly (15%). Amidst Delta in November, total seroprevalence was high (87%) and reached >90% in ≥12-year-olds. Inf-seroprevalence increased further to 26%, with a relative steep rise in the age range from primary school to young adults, peaking around 40%, consistent with a relatively low vaccination coverage. In 2022, total seroprevalence already reached 96% at the beginning of the year and remained high thereafter. Inf-seroprevalence increased sharply to 62% in March after emergence of Omicron-BA.1. Rapid expansion was especially noticeable in <60-year-olds, with estimates around 50% in the youngest ages and up to 90% in adolescents, from where it decreased linearly with age (e.g., 50% in 60-year-olds and 30% in 80-year-olds). After easing of most control measures in spring, overall inf-seroprevalence rose further to 74% in June following Omicron-BA.2. While nearly reaching 100% in adolescents, relative steepest increases were now seen in >40-year-olds (up to 80% in 50-year-olds and 40% in 80-year-olds), despite high booster vaccination uptake earlier. In November, following Omicron-BA.5, inf-seroprevalence increased further to 86% in NL, and >90% among those aged <60 years. Regardless of this relatively large increase in adults during the second half of 2022, still 30% of those vaccinated aged ≥60 years lacked hybrid immunity, and 50% ≥80 years.

Most infections pre-Alpha were observed in the South-East region (15%) and fewest in the North (10%) (**Supplementary Figure S3** displays all 25 Municipality Health Service (GGD) regions). A fairly similar picture was extended across 2021, with steepest increase in the South-East (31%) and Mid-West (27%), and lowest in the North (20%). Low urbanized areas had the highest inf-seroprevalence throughout the study period, but most pronounced in June 2021 following Alpha. Generally, regional differences did not further intensify in 2022. In LVC, inf-seroprevalence was considerably higher in NL, especially noticeable till the end of 2021 (∼1.5 times), and reached >90% after Omicron-BA.5 at the end of 2022. Correspondingly, the total seroprevalence was nearly consistently lower in that period, illustrative of the lower COVID-19 vaccination coverage.

#### (Sub)groups

Both inf- and total seroprevalence did not differ much between sexes. Interestingly, middle-aged women (50-59 years) were significantly more often infected than men already pre-Alpha (16% vs. 9%, respectively, p<0.001) and remained higher throughout 2021-2022 (**Supplementary Figure S4**). Conversely, men aged ≥75 years had on average two times higher inf-seroprevalence up until the beginning of 2022 (39% vs. 24% in women, p<0.001).

Among persons of non-Western descent, inf-seroprevalence rose sharper during Alpha than in Western and native Dutch (∼1.5 times), and this was particularly noticeable among <40-year-olds (**Supplementary Figure S5**). Higher inf-seroprevalence persisted among non-Western in all subsequent waves, yet the total seroprevalence was consistently lowest in this group. Moreover, low/middle-educated persons (from age 25 years) had a significantly higher inf-seroprevalence compared to high-educated already pre-Alpha and throughout 2021 (e.g., June: 21% vs. 17%, p=0.006). This was consistent for all age groups (**Supplementary Figure S6**), but most noticeable in individuals aged ≥60 years (Alpha: 18% vs. 12%, respectively, p<0.001; and amidst Delta: 22% vs. 13%, respectively, p=0.002), coinciding with a lower total seroprevalence in June (p=0.03). Age group-specific seroprevalence estimates became more comparable after the emergence of Omicron. Further, in persons with underlying comorbidities, targeted for early COVID-19 vaccination, a very high total seroprevalence (>95%) was already observed in June 2021. Inf-seroprevalence continued to be relatively low in this group compared to those without up until March 2022 after Omicron BA.1 (45% vs. 70%), yet increased with higher rate thereafter (after Omicron-BA.5: 73% vs. 91%).

#### Occupation

Inf-seroprevalence in the working-age population (18-67 years) followed a similar pattern as the general population but was slightly higher throughout 2021-2022. HCW had among the highest inf-seroprevalence pre-Alpha (20%) – together with transportation and production sectors (**Figure 4AB**) – which was mainly due to high rates in elderly (home)workers (33%) and hospital employees (22%) (**Figure 4CD**). Congruent with a very high total seroprevalence already in June 2021 (>90%) due to prioritization of vaccination, the rate of infections in HCW slowed down (after Alpha: 26%; amidst Delta: 30%) when compared to nationwide, and was similar in March 2022 after Omicron-BA.1. Inf-seroprevalence rates in other HCW sectors were relatively moderate to low until emergence of Omicron-BA.1. Furthermore, sharpest inf-seroprevalence elevations in 2021 were seen among those working in contact professions other than HCW (from 15% in February to 32% in November) and day-care & primary education (from 15% to 29%), and these trends extended into the Omicron waves (e.g., after Omicron-BA.2 (June 2022): 82% and 91%, respectively; to note, the median age was lowest in these sectors: between 43-45 years, vs. others sectors 47-55). Conversely, office employees and those working in middle/higher education had nearly two times lower estimates than HCW after Alpha in June 2021, and only started to catch-up with countrywide estimates thereafter, particularly since emergence of Omicron.

**Figure 4.**
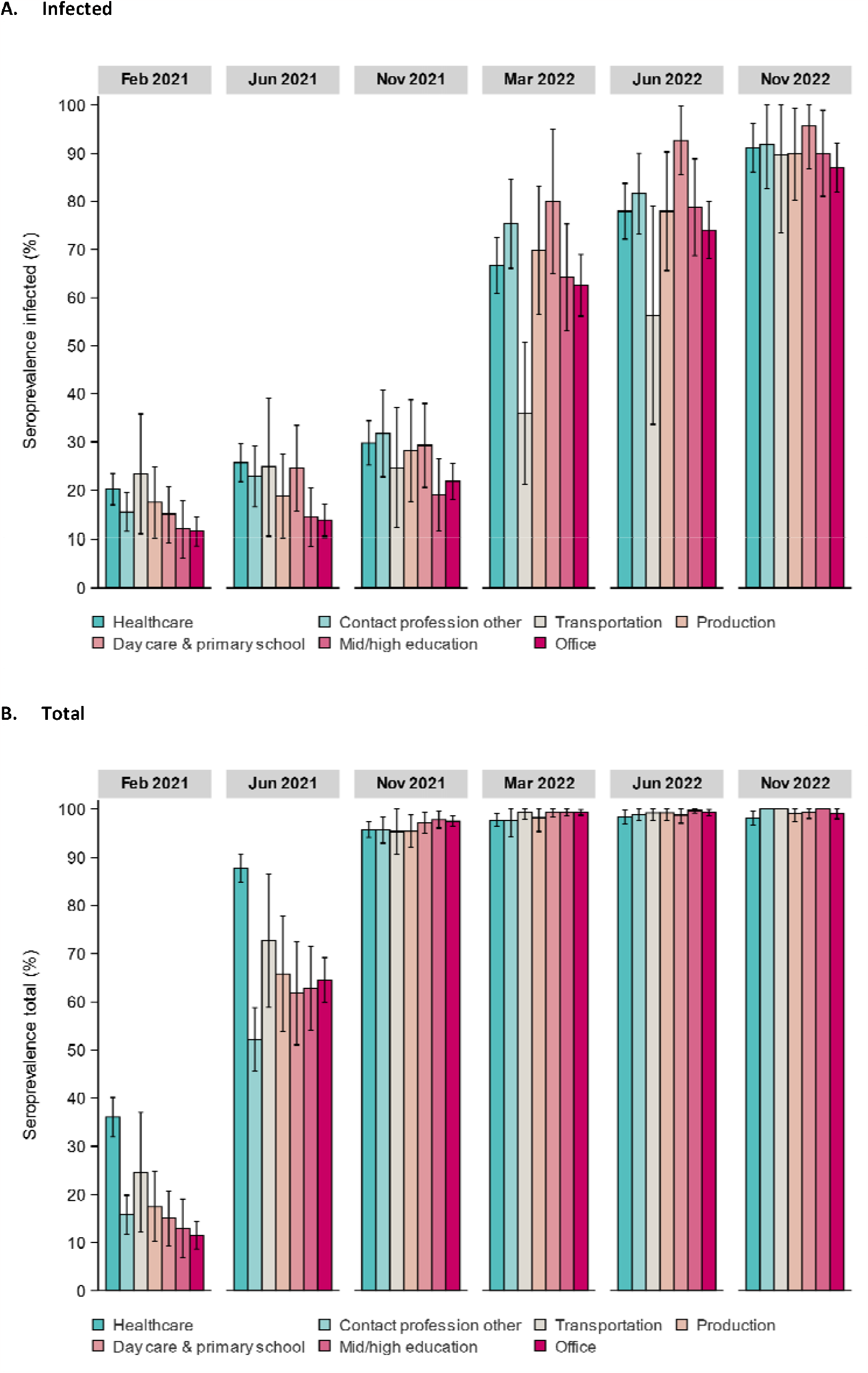
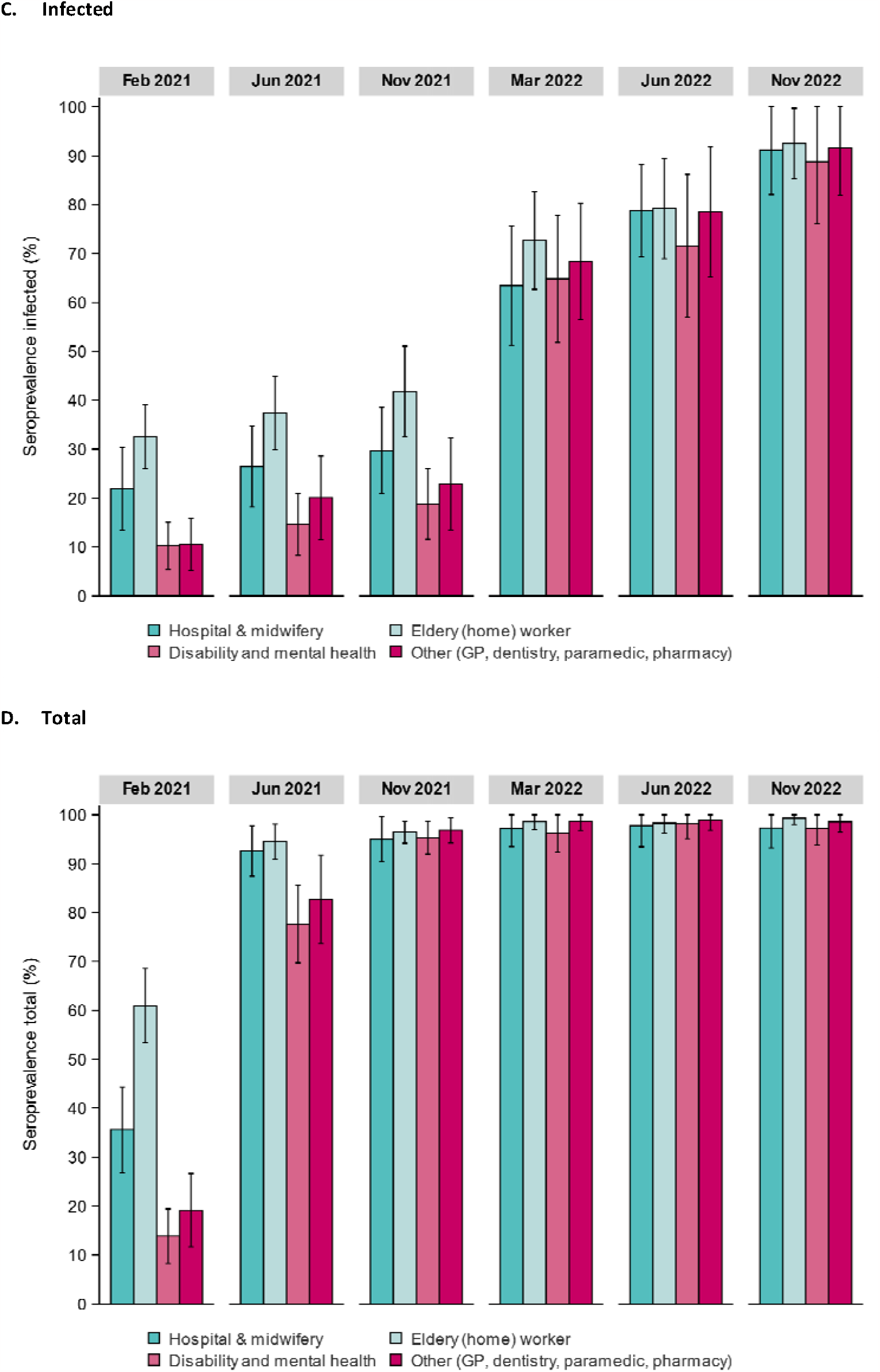
Weighted SARS-CoV-2 seroprevalence (with 95% confidence intervals) induced by infection (A, C) and total (i.e., infection and vaccination) (B, C) in the general Dutch population in 2021 (February (PICO4), June (PICO5) and November (PICO6)) and 2022 (March (PICO7), June (PICO8), November (PICO9)), by age categories (years) and occupational sector (A, B) and healthcare sector (C, D). Contact professions consisted of sectors outside healthcare, such as bars, hotels, restaurants, stores (retail and supermarket), barbers, tattooing, masseuse, cosmetologist, etc. The median age in years (IQR) per occupational sector (in PICO6, as reference) was: Healthcare: 48 (35-57); Contact profession: 43 (22-55); Transportation: 55 (37-60); Production: 49 (37-58); Day care & primary school: 45 (35-56); Middle & higher education: 49 (33-60); and Office: 47 (36-55).

### Breakthrough infections

Primarily in 2021, inf-seroprevalence was significantly lower among adult vaccinees, e.g., amidst Delta in November 2021 (22% vs. 42% among unvaccinated, p<0.001) (**Figure 2**) (notably, in February 2021, inf-seroprevalence was higher among vaccinated due to the high proportion of previously-infected HCW pre-Alpha). This can be explained by presence of only few breakthrough infections detected (February-June 2021, Alpha: 0.5% [95% CI: 0.2-0.7]; and June-November 2021, amidst Delta: 3.5% [2.9-4.3]). Inf-seroprevalence elevated with rather similar rate among vaccinated and unvaccinated since emergence of Omicron-BA.1, and even steeper in vaccinated after Omicron-BA.2 (mid-2022, from 56% to 68%). These data correspond to the large proportion of breakthrough infections observed among vaccinated since Omicron (December 2021-March 2022 (Omicron-BA.1): 43% [40-46]; March-June 2022 (Omicron-BA.2): 23% [21-26]; June-November 2022 (Omicron-BA.5): 28% [25-30]), at first mainly among adolescents up until middle-aged adults, but since Omicron-BA.2 equally across adults up to 80 years (**Figure 5**).

**Figure 5.**
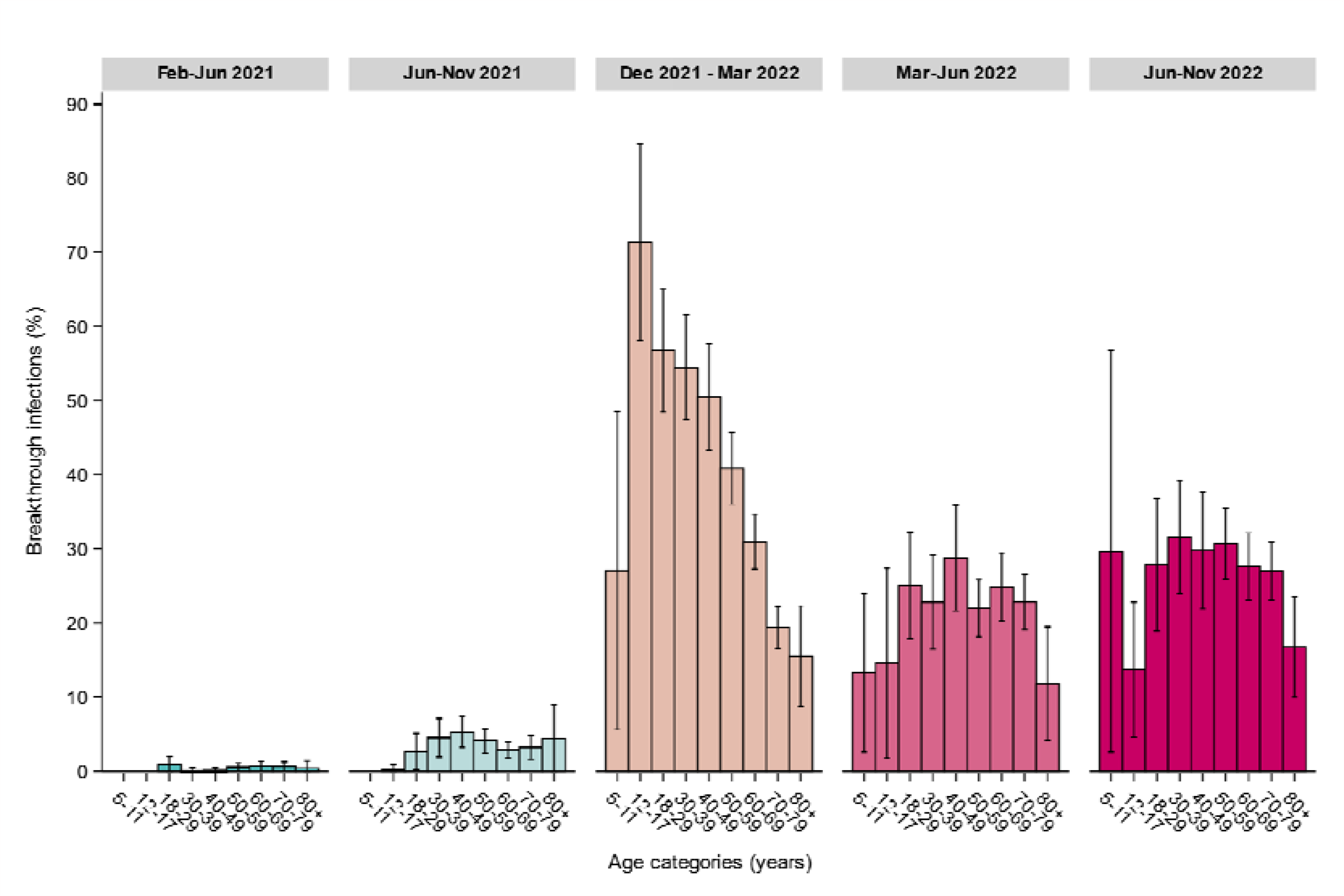
Weighted SARS-CoV-2 breakthrough infections (with 95% confidence intervals) in the general vaccinated Dutch population in 2021 (between February (PICO4), June (PICO5) and November (PICO6)) and 2022 (between March (PICO7), June (PICO8), November (PICO9)), by age categories (years).

## DISCUSSION

This large Dutch nationwide population-based prospective seroepidemiological study supported public health decision-making during the COVID-19 pandemic, e.g., as input for the Outbreak Management Team, Ministry of Health, Dutch Health Council, and forecasting/modelling purposes. Our study underscores the importance of serosurveillance globally to supplement other tools in better understanding population immunity and susceptibility over time. The total seroprevalence in NL increased rapidly during the VOC-era, especially in persons prioritized for vaccination. However, infection rates were unequally distributed between (sub)groups and regions pre-Omicron, with highest rates noticeable in adolescents and young adults, low/middle educated elderly, non-Western, those in contact professions including working in childcare, the South-East region and LVC. Following multiple waves of Omicron, the number of breakthrough infections increased steeply, resulting in hybrid immunity for the majority of the Dutch population by the end of 2022. Nevertheless, approximately three out of ten vaccinated vulnerable individuals (i.e., elderly and those with comorbidities) had not acquired hybrid immunity, necessitating tailored vaccination efforts to reduce the risk of severe disease upon infection and ongoing sero-monitoring.

Our population-based results are indicative of the prevention of infections by vaccination pre-Omicron and in line with vaccine-effectiveness studies [12]. Particular illustrative was the relative modest increase in infections in HCW since vaccination despite having the highest exposure – potentially in combination with adequate use of personal protective equipment. Conversely, the rate of infection remained high in the South-Eastern part of the country, i.e., the epicenter of the first wave in 2020, and particularly in LVC [2]. A combination of lower vaccine uptake (consistent with lower total seroprevalence) and increased exposure, e.g., due to lower adherence to measures, might have caused this and has also been observed in similar communities abroad [4, 22]. Future research should gain more understanding of the concerns in harder-to-reach regions and apply this knowledge as input for tailormade preventive programs. Further, in-depth research on air quality across regions and (the course of) COVID-19 is ongoing, and pre-liminary analysis may point towards a potential increased risk in the lower urbanized areas in the South-East, recognized for poorer air quality due to intensive livestock farming [23].

Importantly, relatively higher rates of infection were also observed in low/middle educated elderly and non-Western, particularly after roll-out of vaccination. The pandemic entails a double burden for the most disadvantaged groups: increased likelihood of infection, e.g., due to fewer opportunities to work remotely, as well as severe disease due to weaker health or access to health. Our data are congruent with lower vaccination uptake and higher ICU admissions in these specific groups in our country and abroad during this period [4, 24, 25]. Global efforts are needed to tackle mis-information and apply tailormade approaches for vaccination, especially since coverage against vaccine-preventable diseases in general is declining.

A distinct age-pattern was seen up until 2021 with infections peaking in adolescents, young adults and middle-aged women. While nationwide stringent control measures were still in effect, primary schools remained open during emergence of VOCs. As opposed to what we observed pre-Alpha [3], the rate of infections in the youngest age groups rose quickly which corresponds to reports on increased transmissibility and susceptibility in children since emergence of VOCs [26]. Consistent herewith, inf-seroprevalence elevated in those working in childcare/primary schools. Pre-Alpha, highest rates of infections were seen in those working in transportation, production and healthcare. Steepest increases were later observed in those working in other contact professions, such as physiotherapists, barbers and store employees, whereas office employees were still among the lowest infected, presumably due to working remotely. Also, despite the highest rates in young adults, professions in middle/higher education were comparable to nationwide estimates. However, unless demonstrative beneficial effects of limited contact and remote schooling on reducing viral transmission, potential detrimental social- and cognitive effects should be taken into account while evaluating the pandemic response [27].

To our knowledge this is the first study to present population estimates of breakthrough infections across various VOCs. After the first Omicron wave and relaxation of control measures, the rate of infections had increased steeply across the population, consistent with reports globally [15], and differences in seroprevalence between groups shrunk. A large elevation in the proportion of breakthrough infections was observed, firstly among adolescents up until middle-aged adults and in later waves across all adults. This trend is most likely explained by increased booster doses in elderly, which have shown to be effective against infection in the short term, but susceptibility increased due to waning immunity alongside novel variants with enhanced immune-escape features [12]. Hybrid immunity has proven to be most effective against future infection [28], potentially mediated by mucosal antibodies in the upper respiratory tract [29], and severe disease [30]. These observations on susceptibility in the most vulnerable persons contributed to targeted prevention strategies with novel booster doses for winter 2023, and warrants ongoing sero-monitoring (of vaccine-effectiveness).

This study has several strengths and limitations that should be highlighted. This is one of the largest prospective population-based serological cohorts, including children and a unique LVC sample, that was set-up since the beginning of the pandemic and is still ongoing. Since (self-)testing has been reduced significantly after relaxation of measures in 2022, serological assessment including boosting of antibodies, is essential for detection of (breakthrough) infections, and for guiding surveillance. Further, we did not observe a significant reduction in sensitivity of anti-N in vaccinees [31], which has been reported by some [32], and although this marker is known to wane quicker than anti-S1 [19, 33], we expect high case ascertainment due to repeated sampling with short intervals. Moreover, in-depth questionnaire data allowed investigation into (sub)groups. Response rates in children were rather low and drop-out rates high, however since children were less restricted by control measures and schools were mostly open, we do not expect large differences between children in terms of exposure, and this was also confirmed by contact data [34, 35]. Despite random selection and weighting our sample, some groups are underrepresented, such as those living in nursing homes – that were hit hard pre-Alpha – and non-Western, who might have refrained from participation due to digital- and/or language barriers. Study participants may generally adhere better to control measures, also reflected by a higher vaccination coverage than the general population, which could have underestimated inf-seroprevalence especially pre-Omicron.

To conclude, this seroepidemiological study has provided important insights on dynamics of SARS-CoV-2 population immunity resulting from (breakthrough) infections and vaccination during the VOC-era. These results will be key in evaluation of control measures, construction of future pandemic response planning, and guiding ongoing preventive strategies for the most vulnerable groups. In the endemic/post-pandemic phase, this robust serological framework will be vital for the relevant challenges ahead, such as unravelling risk factors for post-COVID and mucosal immunity, but also in gaining knowledge on the (potentially permanent) epidemiological changes observed for other respiratory pathogens during the pandemic, e.g., respiratory syncytial virus and group A streptococcal [36, 37].

## STATEMENTS

### Ethical statement

The Medical Research Ethics Committees United (MEC-U) approved the study, conformed to the principles embodied in the Declaration of Helsinki, and all participants provided written informed consent.

## Data Availability

Data are available for researchers upon reasonable request for data sharing to the principal investigator.

## Funding statement

This study was funded by the Ministry of Health, Welfare and Sport (VWS) in the Netherlands.

## Acknowledgements

We greatly acknowledge the participants of the PIENTER Corona (PICO) study. We also wish to thank the logistical research team for all the help concerning organization and logistics, the lab technicians for the serological analyses, and Susan van den Hof for critically reviewing the manuscript.

## Conflict of interest

None declared.

## Supplementary material

### Supplementary information: additional sampling of the national sample, November 2021 (PICO6)

Details on previous sampling up to round 2 of the PICO-study and sample size calculation have been described in Vos *et al*. (2021, JECH) and Vos *et al*. (2021, CID). Due to drop outs in study rounds after PICO2 - particularly notable in younger age groups - and hence to maintain power for subsequent rounds, the Dutch cohort was supplemented with an additional sample of randomly-selected persons from the Dutch population registry (as of October 1^st^, 2021) prior to the start of the 6^th^ study round (November 2021). Congruent with the previous study designs, invitees were randomly drawn from five large regions with roughly the same population size (consisting of provinces: North=Groningen, Friesland, Drenthe and Overijssel; Mid-West=Flevoland and Noord-Holland; Mid-East=Gelderland and Utrecht; South-West=Zuid-Holland and Zeeland; and South-East=Noord-Brabant and Limburg), and from 17 pre-defined age groups (1-4, 5-9, 10-14, 15-19, 20-24, 25-29, 30-34, 35-39, 40-44, 45-49, 50-54, 55-59, 60-64, 65-69, 70-74, 75-79, 80-89 years). A minimum of 400 participants per age stratum was anticipated, taking into account initial response rates as well as drop-out rates per age stratum during the study. Taken together, for the additional sample we randomly selected 66,020 persons, of which 65,690 remained eligible for invitation after initial screening, and of these 3,181 participated. The national sample is further described per study round (4 to 9) in 2021 and 2021 and other relevant characteristics in Table 1 of the main article. Supplementary Table S1 below provides an overview of responders vs. non-responders from the additional sampling, stratified by available sociodemographic variables.

**Supplementary Table S1.**
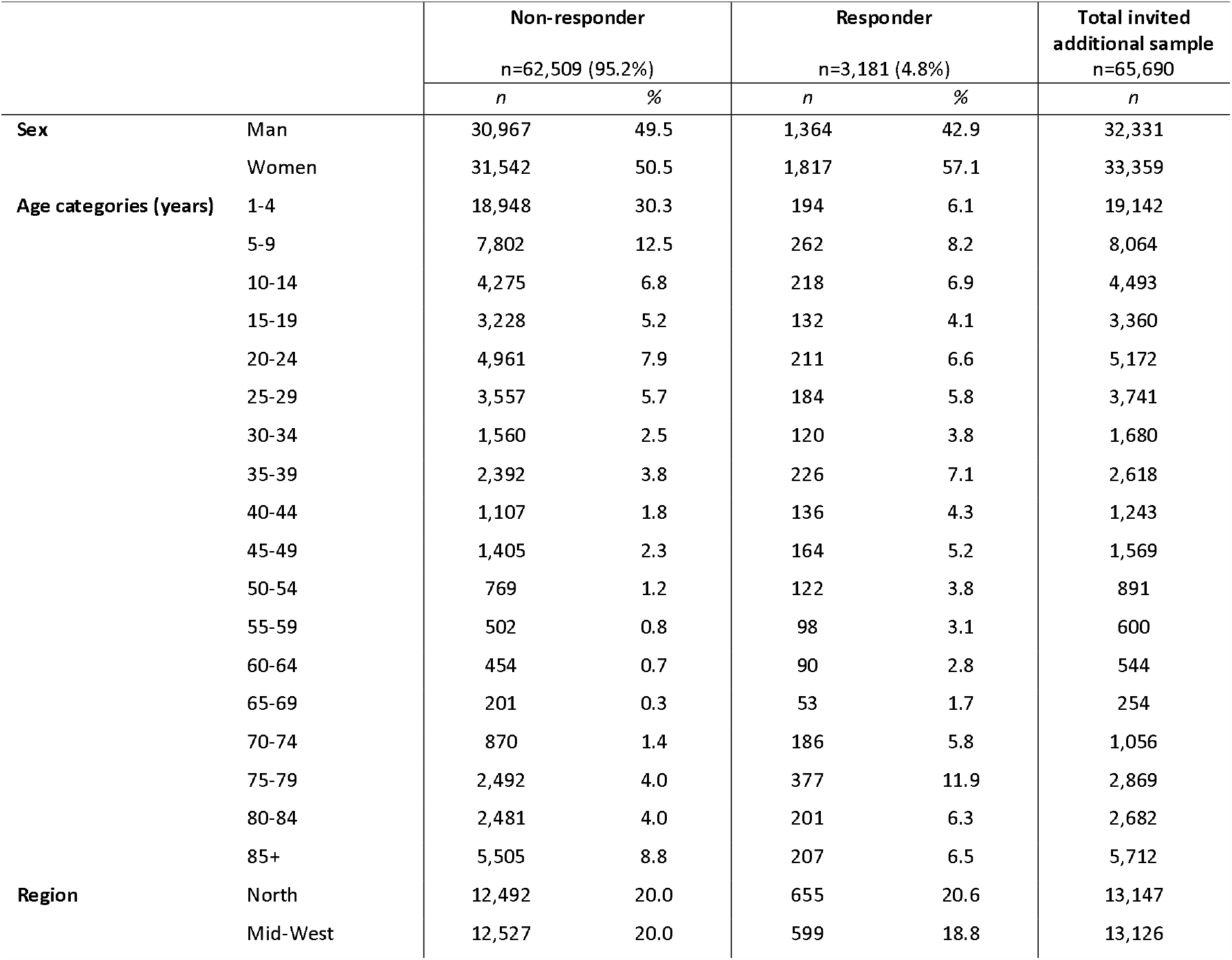
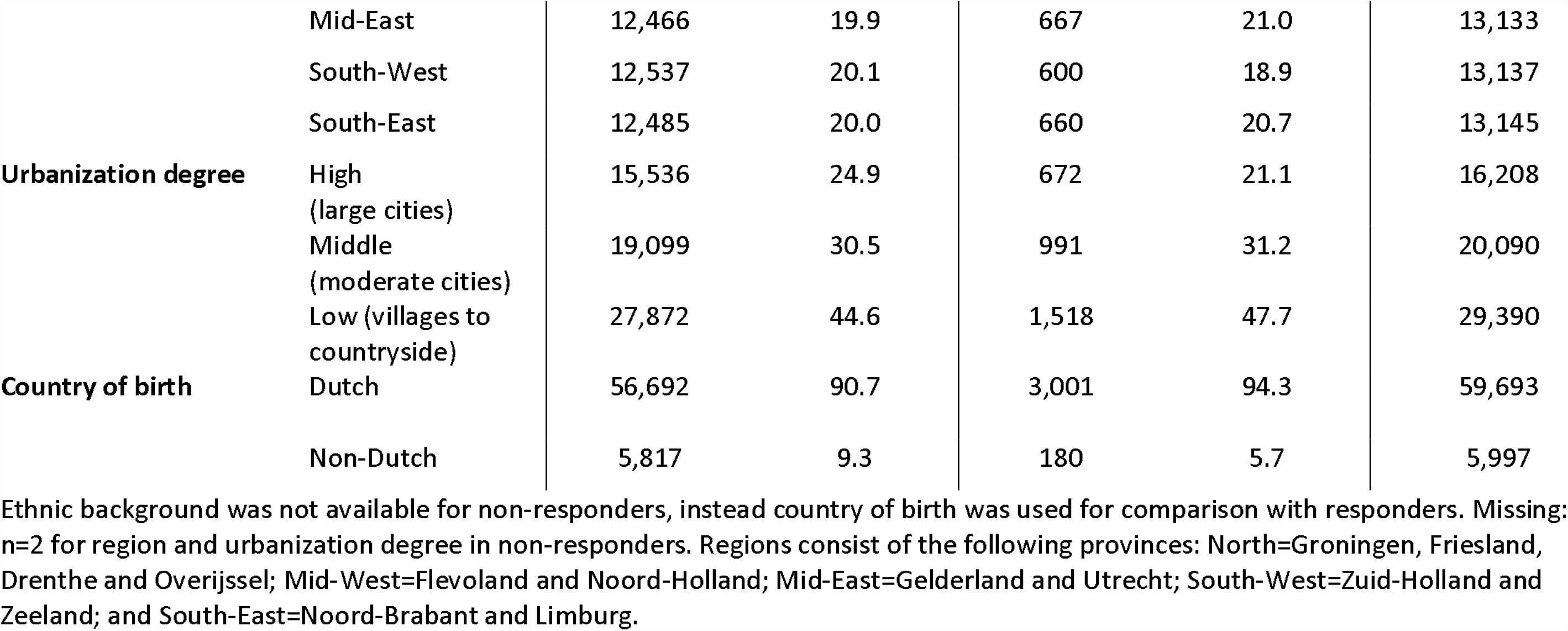
Overview of responders vs. non-responders from the additional sampling in November 2021 (PICO6).

**Supplementary Figure S1.**
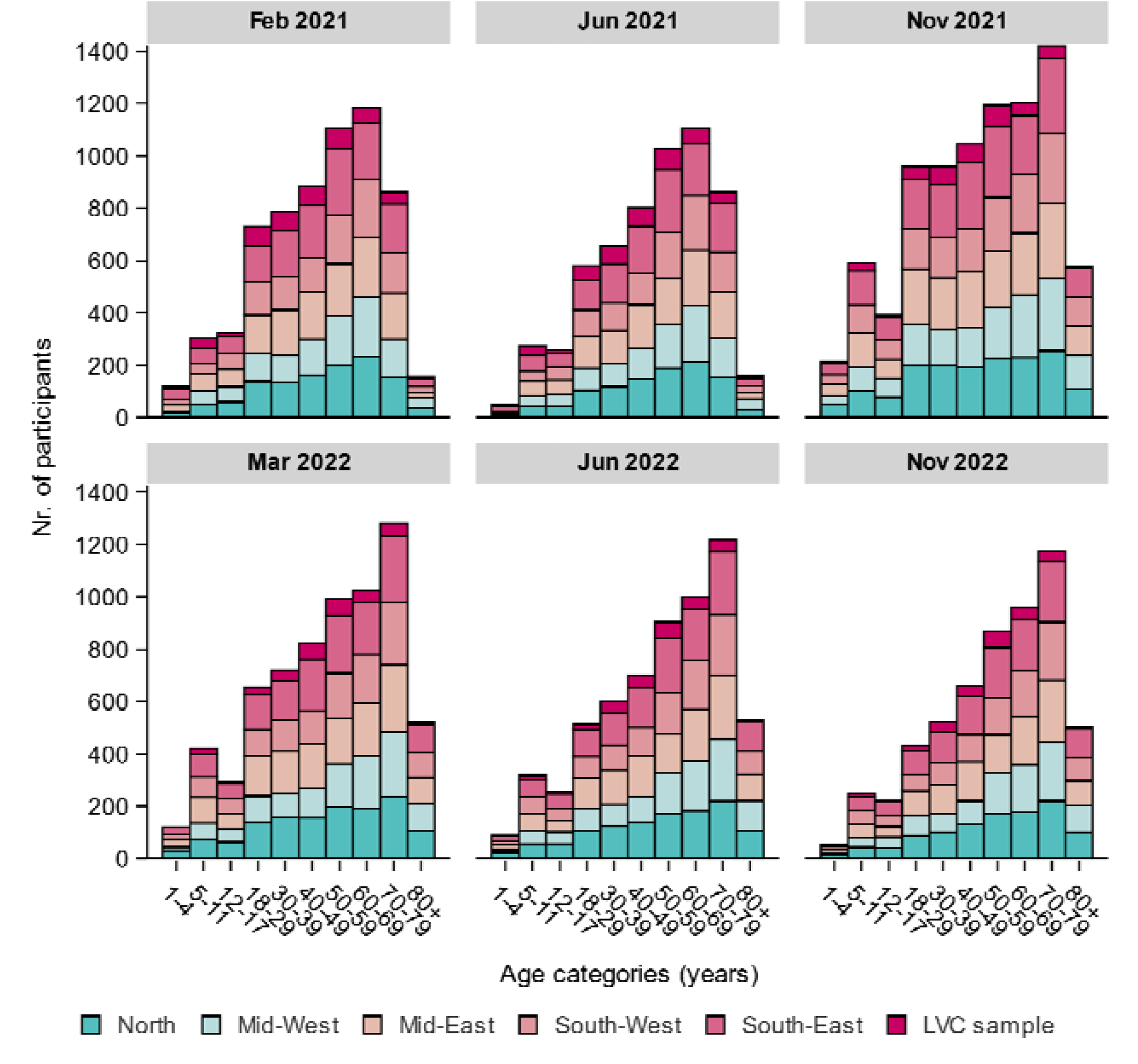
Number of participants in the current PIENTER Corona (PICO) study belonging to the national sample and low vaccination coverage (LVC) sample (as conceived in the PIENTER-3 cohort) in 2021 (February (PICO4), June (PICO5) and November (PICO6)) and 2022 (March (PICO7), June (PICO8), November (PICO9)), by age categories (years) and region. Regions consist of the following provinces: North=Groningen, Friesland, Drenthe and Overijssel; Mid-West=Flevoland and Noord-Holland; Mid-East=Gelderland and Utrecht; South-West=Zuid-Holland and Zeeland; and South-East=Noord-Brabant and Limburg.

**Supplementary Figure S2.**
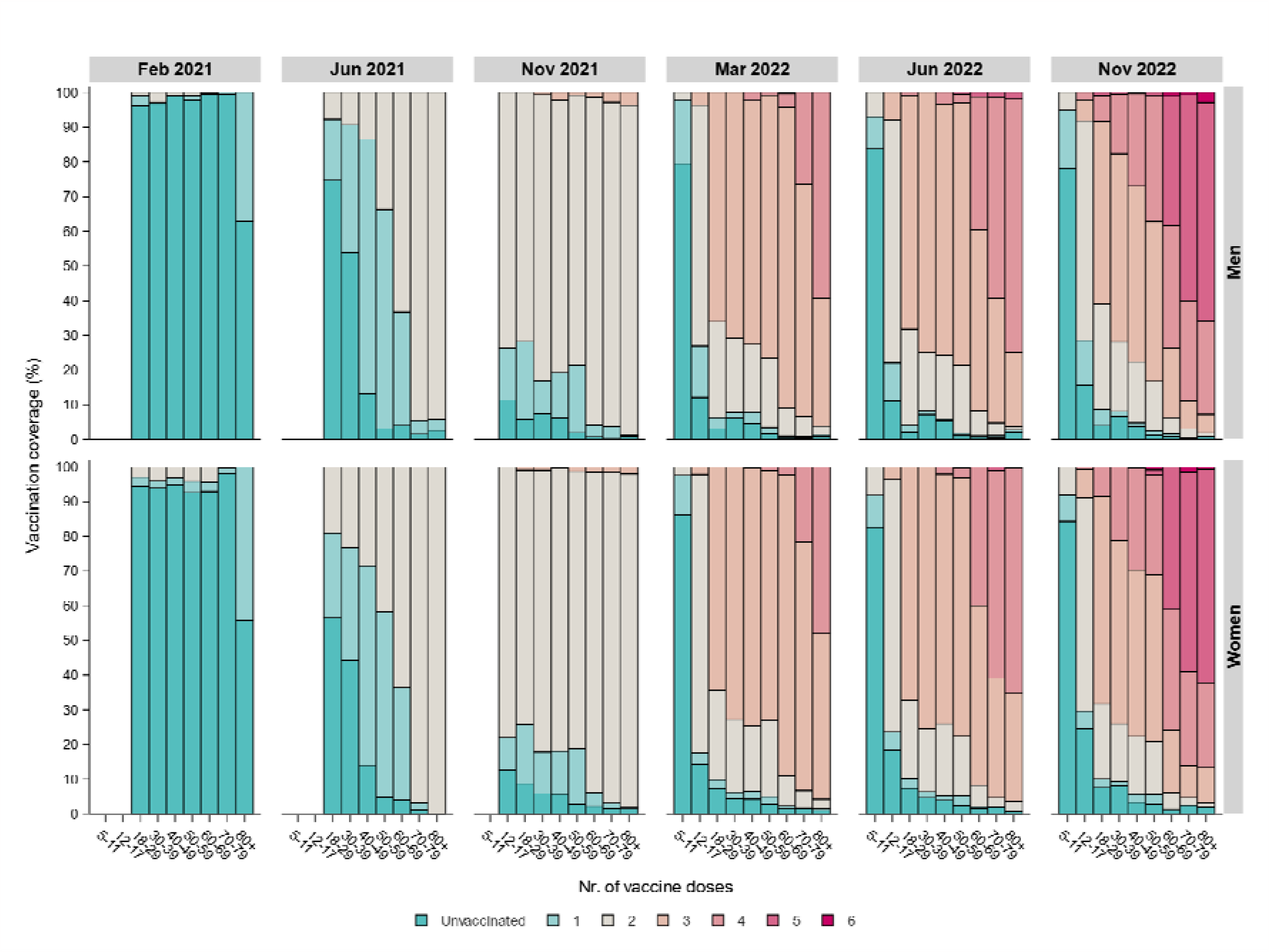
Weighted vaccination coverage (%) in the general Dutch population (as derived from the national sample of the current PIENTER Corona (PICO) study cohort) in 2021 (February (PICO4), June (PICO5) and November (PICO6)) and 2022 (March (PICO7), June (PICO8), November (PICO9)), by sex, age categories (years) and number of vaccine doses. The vaccination campaign roll-out followed an age-, comorbidity-, and healthcare worker-based prioritization that started in the beginning of 2021.

**Supplementary Figure S3.**
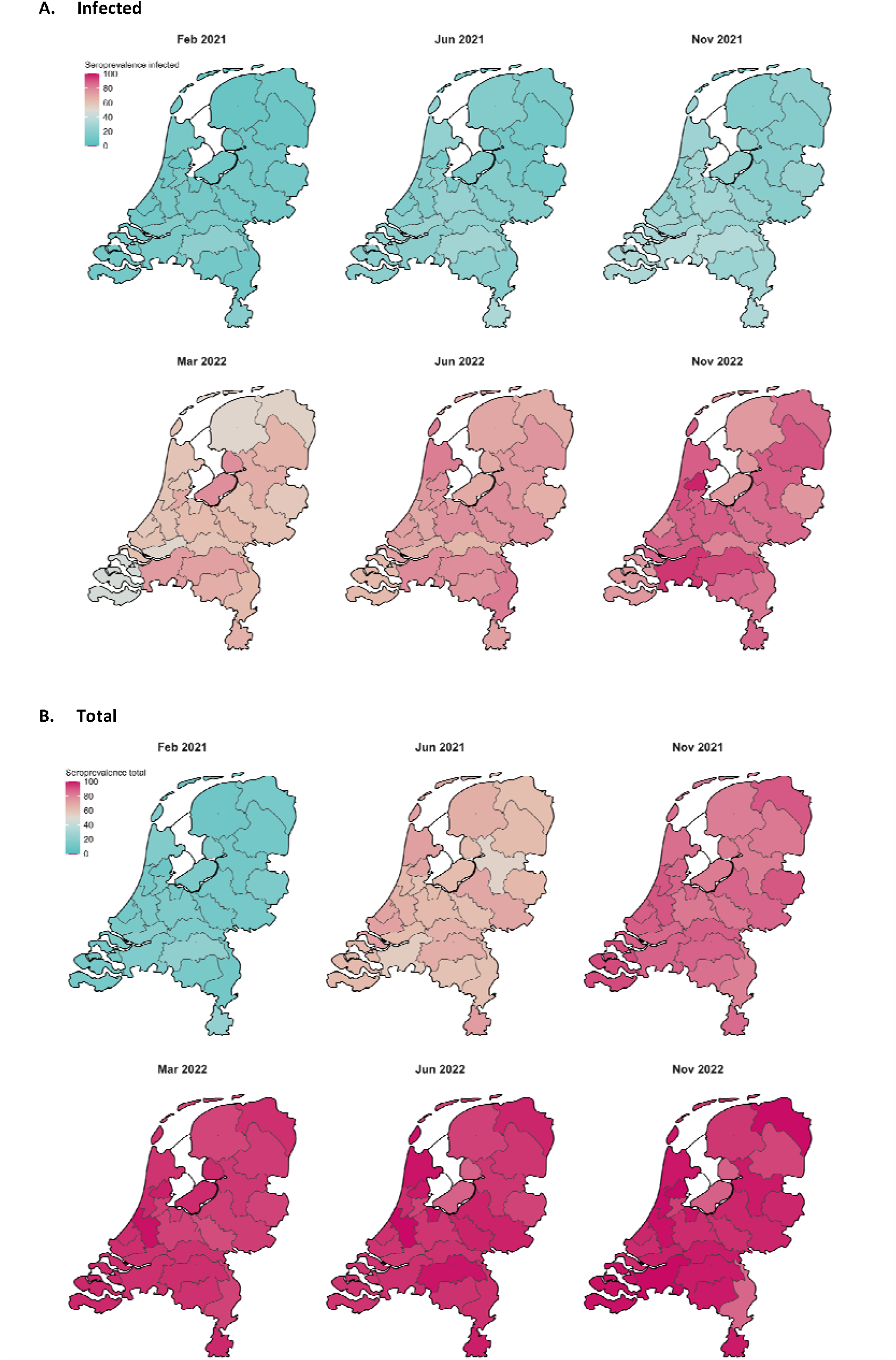
Weighted SARS-CoV-2 seroprevalence induced by infection (A) and total (i.e., infection and vaccination) (B) in the general Dutch population in 2021 (February (PICO4), June (PICO5) and November (PICO6)) and 2022 (March (PICO7), June (PICO8), November (PICO9)), by Municipality Health Service (GGD) region.

**Supplementary Figure S4.**
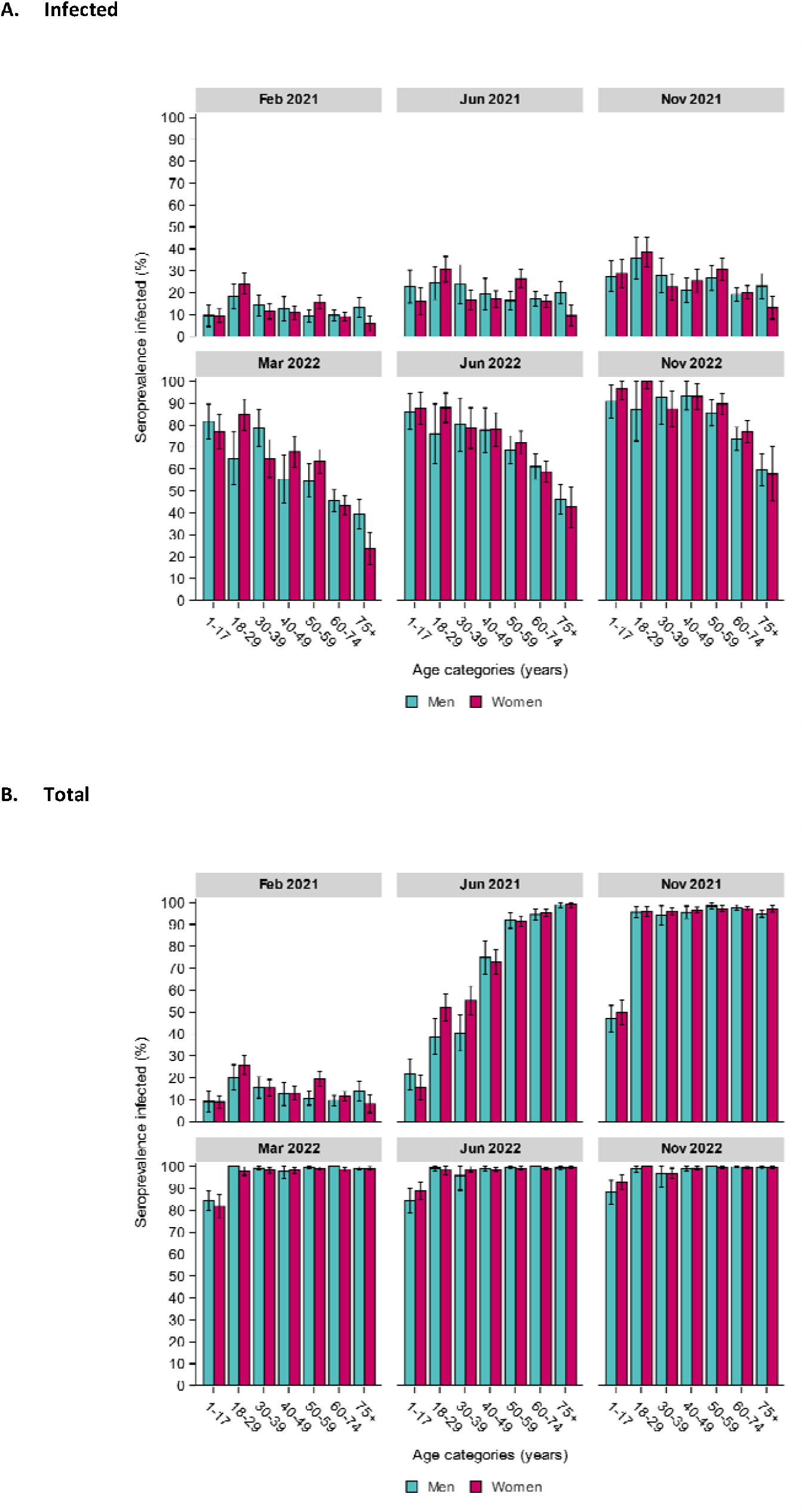
Weighted SARS-CoV-2 seroprevalence (with 95% confidence intervals) induced by infection (A) and total (i.e., infection and vaccination) (B) in the general Dutch population in 2021 (February (PICO4), June (PICO5) and November (PICO6)) and 2022 (March (PICO7), June (PICO8), November (PICO9)), by age categories (years) and sex.

**Supplementary Figure S5.**
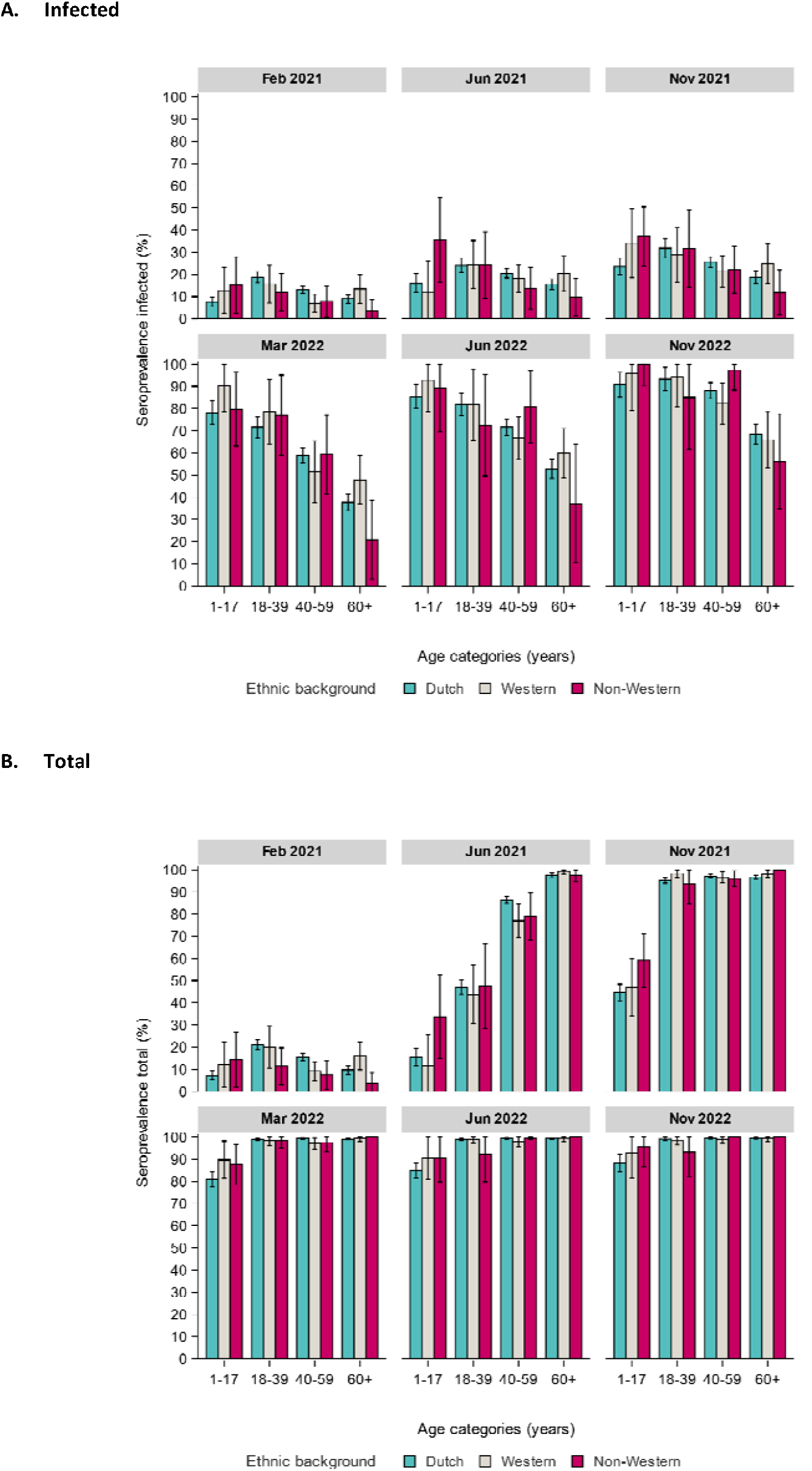
Weighted SARS-CoV-2 seroprevalence (with 95% confidence intervals) induced by infection (A) and total (i.e., infection and vaccination) (B) in the general Dutch population in 2021 (February (PICO4), June (PICO5) and November (PICO6)) and 2022 (March (PICO7), June (PICO8), November (PICO9)), by age categories (years) and ethnic background.

**Supplementary Figure S6.**
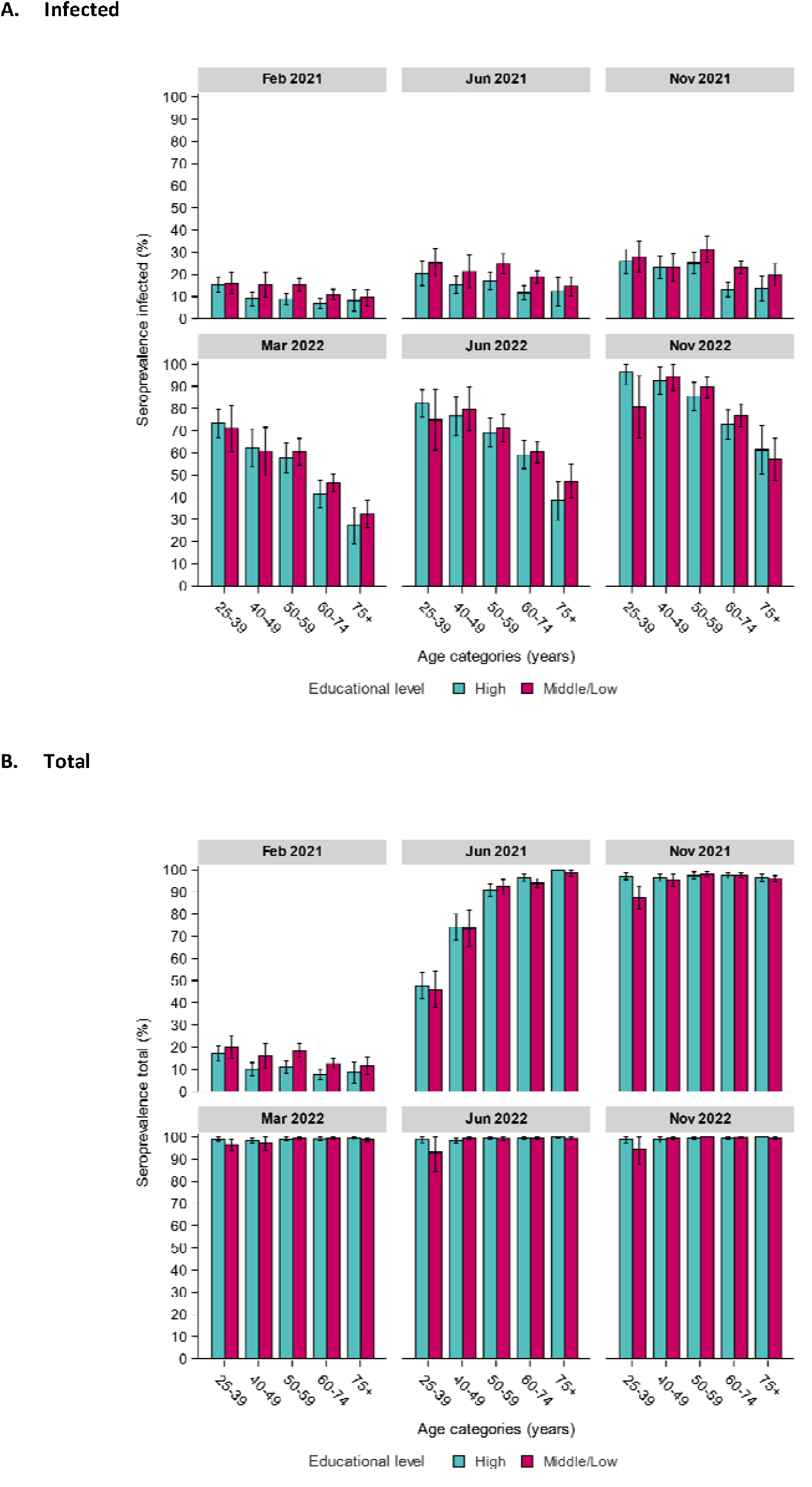
Weighted SARS-CoV-2 seroprevalence (with 95% confidence intervals) induced by infection (A) and total (i.e., infection and vaccination) (B) in the general Dutch population in 2021 (February (PICO4), June (PICO5) and November (PICO6)) and 2022 (March (PICO7), June (PICO8), November (PICO9)), by age categories (years) and educational level ((from 25 years of age, highest obtained or current). Educational level was classified as low (no education or primary education)/middle (secondary school or vocational training), or high (bachelor’s degree, university)).

## Notes

### Competing Interest Statement

The authors have declared no competing interest.

### Author Declarations

The Medical Research Ethics Committees United (MEC-U) in the Netherlands approved the study, conformed to the principles embodied in the Declaration of Helsinki, and all participants provided written informed consent.

